# Preferences for In-Person Consultations with the Referring Physician versus Remote Care provided by the Referring Physician or another Physician among Patients with Long-Term Conditions: A Vignette-Based Survey

**DOI:** 10.1101/2025.06.25.25330282

**Authors:** Tiphaine Lenfant, Elodie Perrodeau, Philippe Ravaud, Viet-Thi Tran

## Abstract

**Objective:** To assess the use of remote care by patients with long-term conditions (LTC) in France to interact with their referring physician (RP), and their preferences for in-person versus remote care with their RP or another physician.

**Methods and Analysis:** Vignette-based survey among adults with at least one LTC recruited from the ComPaRe cohort, a nationwide cohort of patients with LTC in France. Data were weighted to represent French patients with LTC. The survey assessed 1) the availability and use of remote modalities (video consultation, phone contact, and asynchronous message exchange) to interact with their RP, 2) their preferences for in-person consultation or remote modalities to interact with their RP in five situations (worsening symptoms, new symptoms, annual check-up, results discussion, medication renewal), 3) their willingness to trade an in-person consultation in 20 days with their RP for a quicker Direct-to-Consumer (DTC) remote consultation with a non-RP.

**Results:** We included 1995 participants (mean age 55 [SD17], 56% female, 69% with at least 2 LTC). 1) For 47% of patients, remote care with the RP was not available. 2) To interact with their RP, a remote modality was preferred over an in-person consultation by 25% to 55% of patients with higher proportions for results discussion (43%) and medication renewal (55%) than for worsening symptoms (36%), new symptoms (25%), or annual check-ups (26%). 3) Depending on the situation, 20% to 51% of patients were willing to trade an in-person consultation with the RP in 20 days for a DTC remote consultation within 5 days. Factors associated with the preference for a quicker DTC remote consultation were clinical situations (OR 3.16 [2.35-4.26] for new symptoms, p<0.001; OR 3.74 [2.77-5.08] for worsening symptoms, p<0.001, compared to medication renewal), the gender male (OR 1.32 [1.04-1.67], p=0.021), the difficulty to consult in-person (OR 1.54 [1.22-1.96], p<0.001), the unavailability of remote modalities with the RP (OR 1.33 [1.09-1.64], p=0.006), the RP’s specialty (OR 1.79 [1.28-2.50] for general practitioners, p=0.001), medium or poor listening skills of the RP (OR 1.53 [1.20-1.95], p=0.001) and usual delay before the next in-person appointment < 2 weeks (OR 1.43 [1.12-1.82], p=0.003).

**Conclusion:** In France, patients with LTC have limited access to remote care with their RP. Up to 50% of patients would consider replacing an in-person consultation with a remote modality, including with a non-RP, depending on the situation and the delay.

**KEY MESSAGES:** *What is already known on this topic:* Remote care can help reduce the logistical burden of follow-up for patients with long-term conditions (LTC), yet it remains underused in many healthcare systems. Previous studies have shown conflicting findings regarding patient preferences for remote versus in-person care, and few have explored how these preferences vary depending on clinical scenarios or the physician involved.

*What this study adds:* This nationwide, vignette-based survey shows that nearly half of patients with LTCs in France do not have access to remote care with their referring physician. Despite this, up to 55% of patients prefer remote modalities over in-person consultations in some contexts (e.g., medication renewal), and up to 51% are willing to consult an unknown physician remotely if it shortens the delay to care, especially in situations of worsening symptoms or new symptoms.

*How this study might affect research, practice, or policy:* These findings suggest that, in France, remote care is underused not due to patient reluctance but due to limited availability and inadequate integration into care pathways. Policymakers and healthcare providers should prioritize the development of flexible, blended care models that incorporate patient preferences and clinical context to optimize continuity and accessibility of care for patients with LTCs.

## INTRODUCTION

One-third of adults in Western countries have long-term conditions (LTC).^1-3^ The follow-up of LTC, traditionally provided during in-person consultations in current healthcare systems, imposes a logistical and emotional burden on patients.^4^ First, patients must make personal and professional arrangements to attend in-person consultations, with long travel and waiting times. For example, in Germany, a study found that patients needed to allocate over an hour for a 13-minute consultation with a neurologist.^5^ Second, consultations are prescheduled and ill-timed: patients see their physician at a time that is planned in advance, rather than when they need it. For example, guidelines for rheumatoid arthritis recommend consultations every 3 to 6 months, which is neither adapted for the majority of patients who are stable, nor for the minority who will have a flare between visits that still could be missed and left untreated.^6^ Third, between consultations, patients are left alone to manage their conditions, with limited guidance on when and how to seek help.^7^ All these burdens are amplified for patients with multiple LTC, who represent approximately 50% of patients with LTC, and who must deal with these issues across all their different providers.^1-3,8,9^

Remote care could address some of these limitations by reducing travel time and facilitating communication at the right moment.^4,10,11^ Remote care can be either synchronous (via video consultations or phone contacts) or asynchronous (via secure messaging on secure health platforms or patient portals). Synchronous modalities mimic the dynamics of an in-person consultation by enabling real-time interactions between the patient and the physician, while alleviating geographical and personal constraints.^12,13^ Asynchronous modalities enable patients to send information, ask questions, or share documents with their physicians at any time, without the fear of disturbing and waiting for availability, as responses are not expected in real time.^14,15^ Beyond communication modalities, remote care can be delivered by the referring physician or by a non-referring physician through Direct-to-Consumer (DTC) remote consultations.^16^

In the literature, studies provide conflicting results on patients’ preferences about remote care. On one hand, when asked about a single consultation, patients often prefer in-person care, which allows for human contact and physical examination.^13,17-19^ On the other hand, when asked about how they envision their long-term management, 31% of patients imagine that video consultations could replace in-person contacts for more than 50% of their consultations.^10^ Moreover, few studies have investigated preferences for remote care based on specific clinical contexts, such as stable versus unstable long-term conditions, medication renewals, or discussions of results.^10,15,20^ Finally, it is unclear whether patients would be willing to use DTC remote consultations when their referring physician is unavailable within an acceptable timeframe.^16^

We thus aimed to evaluate, among patients with LTC in France: 1) the availability and use of three remote modalities (video consultation, phone contact, and asynchronous message exchange) to interact with their referring physician (RP), 2) their preferences between an in-person consultation and these three remote modalities to interact with their RP in five situations (worsening symptoms, new symptoms, annual check-up, results discussion, or medication renewal), 3) their willingness to trade an in-person consultation in 20 days with their RP for a quicker remote consultation with a non-RP in these situations.

## METHODS

We conducted an online vignette-based survey to assess patients’ preferences regarding the follow-up of their LTC. Vignettes are descriptions of hypothetical scenarios followed by questions to elicit the participants’ preferences.^21,22^ Vignette-based surveys have shown to adequately represent real-world behaviors, including in situations with high desirability bias.^21,22^ For this online survey, we used the Checklist for Reporting Results of Internet E-Surveys (CHERRIES) reporting guideline when applicable.^23^

### Participants

We recruited adults (≥18 years of age) with any long-term condition (LTC), defined as a condition requiring healthcare for more than six months, from ComPaRe, a nationwide e-cohort of more than 60 000 patients with LTC in France (as of May 2025) who donate time to participate in research.^24^ Patients who had been recently active in the e-cohort (i.e., who had logged on to their ComPaRe account within the 12 months preceding the study) and who had consented to be invited to external studies were invited to participate. ComPaRe was approved by the Institutional Review Board of Hôtel-Dieu Hospital in France. Patients provided informed consent. The study protocol received ethical approval from CER Université Paris Cité on October 4th, 2023 (IRB 00012023-85).

### Survey Design and Administration

Participants were invited to complete an online survey, which was structured into three parts. First, the survey assessed the availability and use of three remote modalities (video consultation, phone contact, and asynchronous message exchange) with their RP. It also gathered demographics, health data, and information about their RP (***Suppl Text 1***).

Second, the survey assessed patients’ preferences between an in-person consultation and the three remote modalities to interact with their RP, by presenting each participant with one randomly assigned situation (vignette) among five. Each vignette started with the prompt *“Imagine the following situation:”* and was followed by a description of the situation:

> *Vignette 1: “You are experiencing a worsening of your usual symptoms. You know these symptoms because you have had them in the past*.*”*
>
> *Vignette 2: “You have a new symptom. It is a symptom you have never had before. You do not think it requires an emergency visit*.*”*.
>
> *Vignette 3: “You need an annual check-up on your long-term conditions. Your symptoms are stable*.”
>
> *Vignette 4: “You need to renew the prescription for your long-term treatment. Your symptoms are stable*.*”*
>
> *Vignette 5: “You have received test results for your long-term condition(s). You have not noticed anything alarming in the results*.*”*

The vignettes ended with “*You would like to consult your referring physician*.” Patients were presented with four options: a traditional in-person consultation (IP), a video consultation (VC), a phone contact (PC), or an asynchronous message exchange (AME) via a secure online platform.

Third, the survey assessed patients’ willingness to trade an in-person consultation in 20 days with their RP for a quicker remote consultation with a non-RP in the same situation (same vignette), by asking, *“In this situation, imagine you could see your referring physician in person in 20 days, would you rather see an unknown physician that could see you remotely sooner?”*. Options were

1. *“Yes, I would like to see this unknown physician remotely sooner*.*”*
2. *“It depends on the delay to this remote consultation with the unknown physician*.*”*
3. *“No, I would rather see my referring physician in person in 20 days*.*”*

If they opted for *“It depends on the delay”* (option 2), they were provided with four successive delay reductions for the DTC remote consultation: “*What if the remote appointment with this unknown physician was in 15 to 20 days*”, “*in 10 to 15 days*”, “*in 5 to 10 days*”, or “*within 5 days*”, and could opt for the DTC remote consultation with the non-RP in this delay or wait for the in-person consultation with their RP in 20 days.

The survey was tested in a pilot phase of 30 patients with LTC, recruited through personal contacts. Each patient completed the survey and reviewed each question with the principal investigator (TL) to ensure its clarity, reproducibility, and reliability, as well as the site’s functionality and ease of use. The pilot phase led to iterative refinements of the question wording and response options. At the end of the pilot phase, all 30 patients reviewed the final version of the survey. The survey was available online on PROCESS (Platform for Research Online and CitizEn Science Surveys), accessible from a computer or any other connected device (tablet, smartphone) via a link received by email. Participation was anonymous.

### Statistical analysis

To obtain estimates representative of the French population of patients with LTC, we performed analyses on a weighted data set with calibration on margin with weights for gender, age categories (18-24, 25–34, 35–44, 45–54, 55–64, 65–74, and >75 years), and educational level from the 2021 survey of Santé Publique France.^25^ Descriptive results are presented on the weighted data set. Analyses were conducted using R (version 4.3.0).

First, we described the availability and use of the three remote modalities (VC, PC, AME) with the RP.

Second, we described the proportion of patients preferring a remote modality over an in-person consultation with their RP in each of the five situations. We analyzed the three modalities of remote care (VC, PC, AME) separately and then pooled them into a single “remote” variable. We conducted a logistic regression model to identify the factors associated with patient preferences for remote care. The dependent variable was the preference, pooled into a binary variable (in-person *versus* remote). Covariates were the characteristics of patients, physicians, and consultations (***Suppl Text 2***). We also targeted the subgroup of patients who had never experienced any of the three remote modalities to interact with their RP.

Third, we described the proportion of patients willing to trade an in-person consultation with their RP for a quicker DTC remote consultation with a non-RP, in each of the five situations, for each delay reduction. We conducted a logistic regression model to identify the factors associated with this preference. Dependent was the preference pooled in a binary variable: “No” (answer 3 *“I would rather see my referring physician in person in 20 days*”) *versus* “Yes or Open” (answers 1 and 2: *“It depends on the delay,”* or *“Yes, I would like to see this unknown physician remotely sooner*.*”*). Covariates were the characteristics of patients, physicians, and consultations (***Suppl Text 2***).

## RESULTS

From April 1^st^ to August 28^th,^ 2024, 2328 patients consented to participate in the study, 2038 completed the study, and 1995 had sufficient data to compute the calibration of margin and were analyzed (weighted population), see Flow chart (***Suppl Figure 1***).

After weighting, the mean age was 55 years, 56% of patients were female, and 69% were multimorbid. The most reported conditions were rheumatologic diseases (31%), high blood pressure (30%), mental health diseases (27%), dermatologic diseases (16%), and hepatic/gastroenterological diseases (15%); diabetes affected 15% of patients, and 9% had cancer or hematologic diseases. Participants’ RP (i.e., the physician who provided the most regular follow-up for their LTC) was a general practitioner in 68% of cases. The delay between departure from home or work and the start of an in-person consultation was over an hour for 37% of patients. In total, 79% of patients reported it easy to free up time (half a day or more) for an in-person consultation (***Table 1 and Suppl Table 1***).

**Table 1.** Patients’, physicians’, and consultations’ characteristics.

|  | Raw data set<br>N = 1995 | Weighted data set<br>N = 1995 |
| --- | --- | --- |
| **Patients' characteristics** |  |  |
| **Age — years (mean, SD)** | 52 (15) | 55 (17) |
| **Gender — no. (%)** |  |  |
| Male | 562 (28%) | 882 (44%) |
| Female | 1433 (72%) | 1113 (56%) |
| **Highest diploma - no. (%)** |  |  |
| Higher than a bachelor's degree | 1209 (61%) | 345 (17%) |
| Bachelor's degree or lower | 786 (39%) | 1650 (83%) |
| **Self-perception of financial situation — no. (%)** |  |  |
| Difficult | 697 (35%) | 771 (39%) |
| Not difficult | 1298 (65%) | 1224 (61%) |
| **Multimorbidity (≥2 LTC) — no. (%)** | 1323 (66%) | 1384 (69%) |
| **Self-assessed anxiety level (score from 1 to 6, 6 being the highest level of anxiety) — no. (%)** |  |  |
| ≤ 4 / 6 | 1201 (60%) | 1168 (59%) |
| > 4 / 6 | 794 (40%) | 827 (41%) |
| **Self-management (score from 1 to 6, 6 being the best self-management) — no. (%)** |  |  |
| Good to Excellent | 1248 (63%) | 1183 (59%) |
| Poor to Medium | 747 (37%) | 812 (41%) |
| **Digital Health Care Literacy Scale (score from 3 to 15, 15 being the highest digital literacy) — no. (%)** |  |  |
| Higher DHCLS (>10 / 15) | 1729 (87%) | 1646 (83%) |
| Lower DHCLS (≤10 / 15) | 266 (13%) | 349 (17%) |
| **Reported Long-term conditions (LTC)** |  |  |
| Rheumatologic diseases | 558 (28%) | 609 (31%) |
| Mental health diseases | 530 (27%) | 533 (27%) |
| High blood pressure | 439 (22%) | 591 (30%) |
| Gynecological diseases | 373 (19%) | 231 (12%) |
| Endocrine diseases | 295 (15%) | 280 (14%) |
| Hepatic and gastroenterological diseases | 294 (15%) | 301 (15%) |
| Dermatologic diseases | 277 (14%) | 313 (16%) |
| Inflammatory and Immune-mediated diseases | 233 (12%) | 233 (12%) |
| Neurologic diseases | 229 (12%) | 250 (13%) |
| Pulmonary diseases | 221 (11%) | 249 (13%) |
| Diabetes | 203 (10%) | 295 (15%) |
| Ophthalmologic diseases | 193 (10%) | 239 (12%) |
| Cardiac diseases | 179 (9%) | 247 (12%) |
| Hematologic and oncologic diseases | 161 (8%) | 184 (9%) |
| Kidney diseases | 108 (5%) | 125 (6%) |
| Infectious diseases | 46 (2%) | 41 (2%) |
| Other | 525 (26%) | 513 (26%) |
| **Referring Physician's characteristics** |  |  |
| **Physician's specialty — no. (%)** |  |  |
| General Practitioner | 1259 (63%) | 1356 (68%) |
| Specialist | 735 (37%) | 639 (32%) |
| Missing | 1 (0%) | 0 (0%) |
| **Length of relationship with the physician — no. (%)** |  |  |
| 5 Years Or More | 1167 (58%) | 1176 (59%) |
| Less Than 5 Years | 821 (41%) | 811 (41%) |
| Missing | 7 (0%) | 8 (0%) |
| **Listening skills evaluated by the patient — no. (%)** |  |  |
| Good to Excellent | 1553 (78%) | 1533 (77%) |
| Poor to Medium | 440 (22%) | 462 (23%) |
| Missing | 2 (0%) | 0 (0%) |
| **Consultations' characteristics** |  |  |
| **Time from departure to the start of an in-person consultation (transportation, parking, inscription, waiting room) — no. (%)** |  |  |
| < 1 Hour | 1195 (60%) | 1244 (63%) |
| ≥ 1 Hour | 795 (40%) | 743 (37%) |
| Missing | 5 (0%) | 8 (0%) |
| **Delay before the next available in-person consultation — no. (%)** |  |  |
| < 2 Weeks | 1018 (51%) | 1121 (56%) |
| ≥ 2 Weeks | 970 (49%) | 865 (44%) |
| Missing | 7 (0%) | 9 (0%) |
| **Ability to make personal/professional arrangements to free up time to attend an in-person consultation — no. (%)** |  |  |
| Difficult to Impossible | 564 (28%) | 419 (21%) |
| Ability to free up half a day or the entire day | 1429 (72%) | 1575 (79%) |
| Missing | 2 (0%) | 1 (0%) |
| **Most frequently used modality — no. (%)** |  |  |
| In-person consultation | 1856 (93%) | 1875 (94%) |
| Phone contact | 12 (1%) | 8 (0%) |
| Video consultation | 102 (5%) | 92 (5%) |
| Asynchronous message exchange | 24 (1%) | 20 (1%) |
| Missing | 4 (0%) | 0 (0%) |
| **Availability and Use of Remote modalities — no. (%)** |  |  |
| Not available or never used (unserved) | 857 (43%) | 943 (47%) |
| Used | 1137 (57%) | 1052 (53%) |
| Missing | 1 (0%) | 0 (0%) |

### Availability and use of remote modalities to interact with the referring physician

In total, data from 1994 (99.9%) patients were available to describe the availability and use of remote modalities to interact with the referring physician. To interact with the RP, video consultations were available to 28% of patients, phone contacts to 11%, and asynchronous message exchanges to 32% (***Figure 1***). For 47% of patients, none of the three remote modalities (VC, PC, AME) was available (unserved patients).

**Figure 1.**
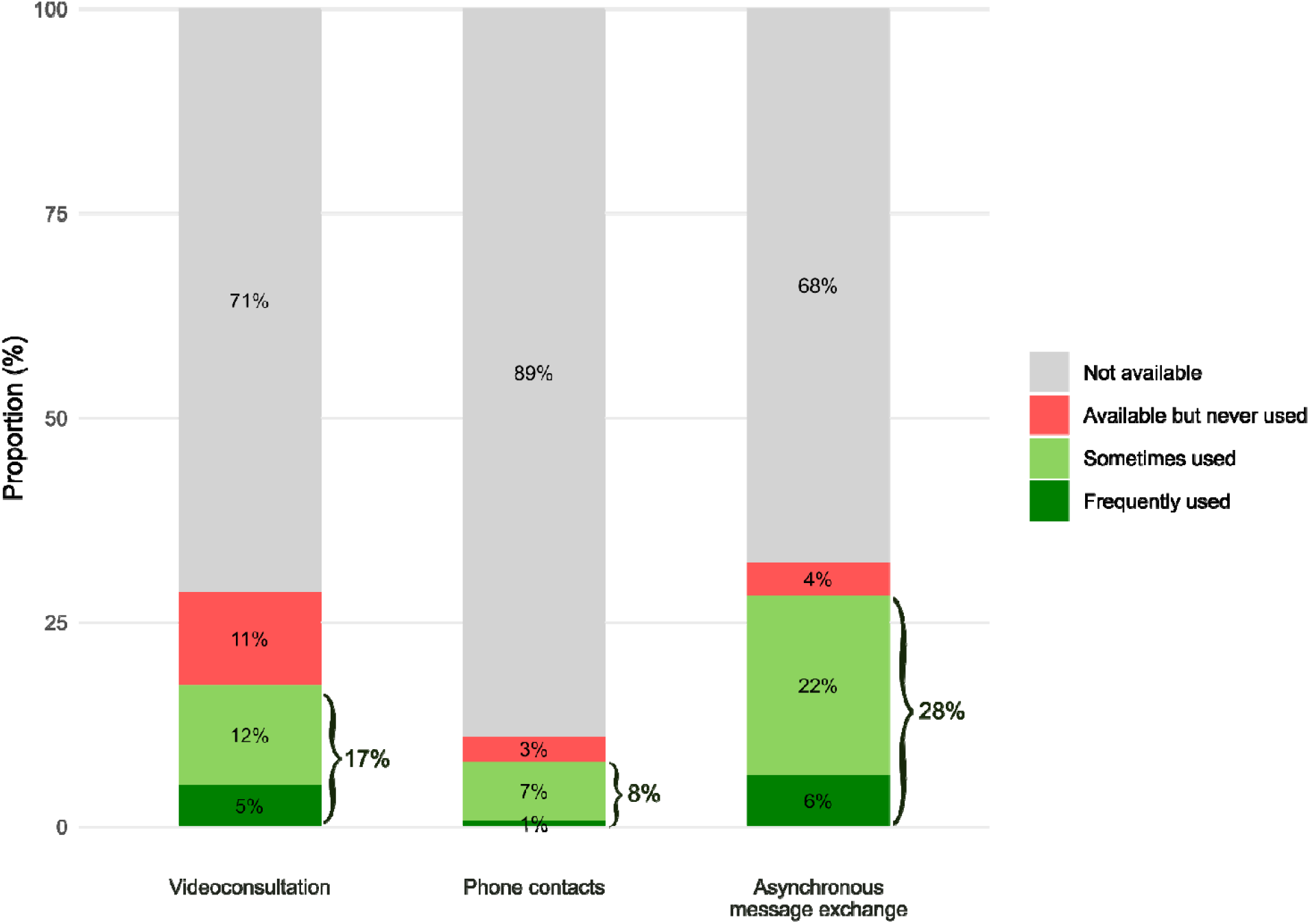
Availability and use of remote modalities to interact with the referring physician (weighted data set, n=1994) Legend: The referring physician is identified by the patient the physician providing the most regular follow-up of their long-term condition(s).

Patients’ preferences for in-person consultations or remote modalities with their RP, depending on the situation

In total, data from 1908 (96%) patients were available to describe their preferences for in-person consultations or remote modalities with their RP. Across the five situations, a remote modality was preferred by 37% of patients. This preference for remote care was higher for results discussion and medication renewal (43% and 55%, respectively) than for worsening symptoms, new symptoms, and annual check-ups (36%, 25%, and 26%, respectively) (***Figure 2***).

**Figure 2.**
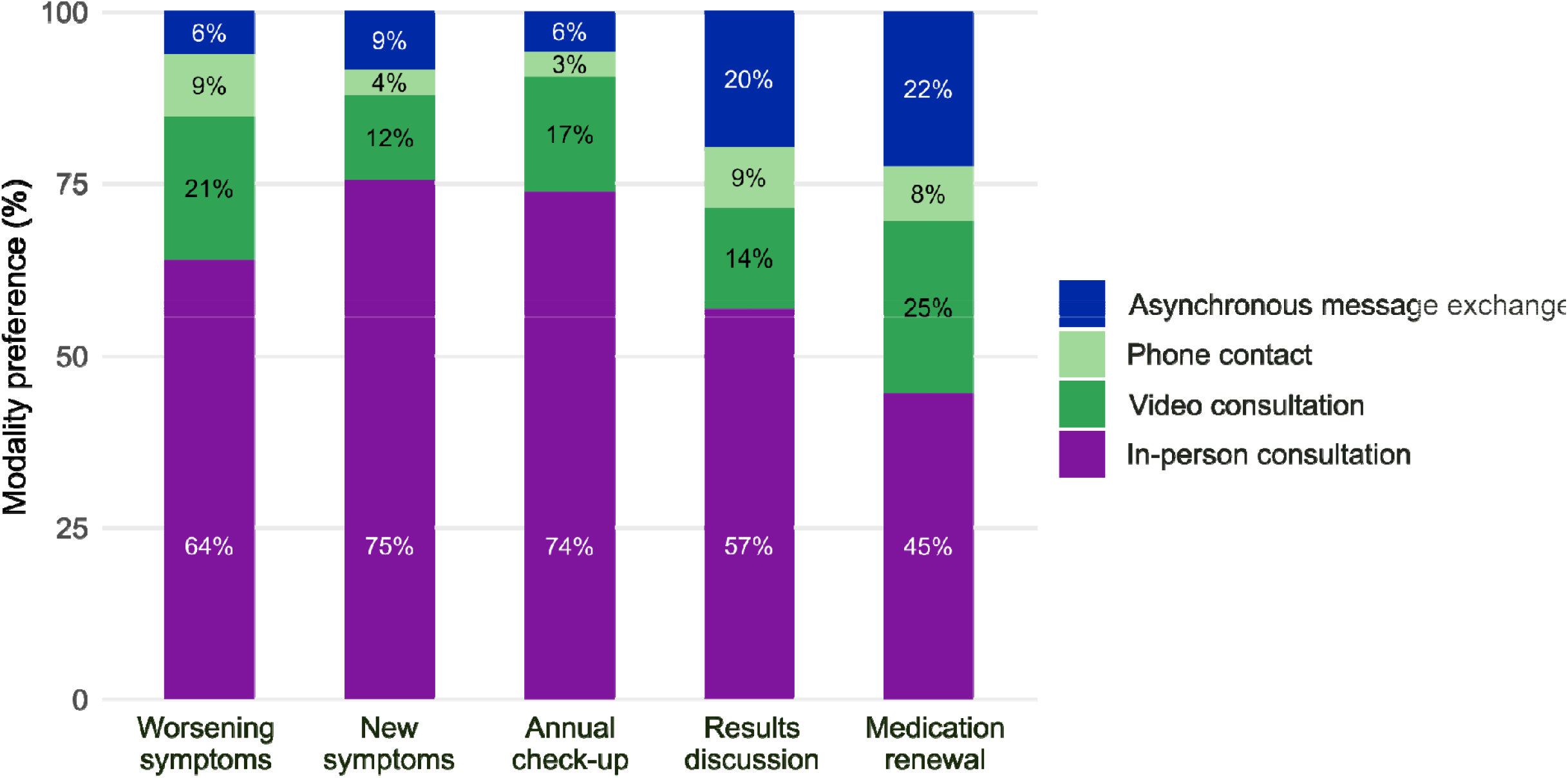
Patients’ preferences for in-person consultation or remote modalities to interact with their referring physician in five clinical situations (weighted data set, n=1908) Legend: The referring physician is identified by the patients as the physician providing the most regular follow-up of their long-term condition(s).

These results were similar among the 47% of unserved patients, with 36% of them preferring an in-person consultation in the situation they were assigned. This preference varied from 18% for new symptoms to 54% for medication renewal (***Suppl Figure 2***).

In the logistic regression model (***Suppl Table 2***), being unserved was not associated with the preference for in-person consultations over remote modalities (OR 1.06 [0.86-1.30], p=0.582). Three clinical situations were associated with the preference for an in-person consultation: worsening symptoms (OR 0.39, 95%CI [0.29-0.53], p<0.001), new symptoms (OR 0.28, 95%CI [0.21-0.38], p<0.001), and annual check-up (OR 0.27, 95%CI [0.20-0.37], p<0.001), with medication renewal as the situation reference. Patients’ characteristics such as male gender (OR 0.73 [0.57-0.92.], p=0.008), lower digital health literacy (OR 0.51 [0.37-0.70], p<0.001) and the easiness to consult in-person (OR 0.47 [0.37-0.60], p<0.001) were associated with the preference for an in-person consultation. Physicians’ characteristics such as medium or poor listening skills (OR 1.35 [1.06-1.73], p=0.016) and longer delays before the next in-person appointments (OR 1.38 [1.09-1.74], p=0.007) were associated with the preference for a remote modality. The subgroup analysis among unserved patients found similar results (***Suppl Table 3***).

### Patients’ willingness to trade an in-person consultation with their RP for a quicker Direct-to-Consumer remote consultation with a non-RP

In total, data from 1940 (97%) patients were available to describe their willingness to trade an in-person consultation with their RP in 20 days for a quicker Direct-to-Consumer remote consultation with a non-RP (***Suppl Table 4***). This willingness depended on the situation and increased with the reduction in delay (***Figure 3***). Patients were more willing to trade for a quicker DTC remote consultation for worsening symptoms or new symptoms than for annual check-ups, medication renewal, or results discussion. When confronted with worsening symptoms, 51% of patients preferred a DTC remote consultation within 5 days rather than waiting 20 days to consult their RP in person. In contrast, for their annual check-up, 20% of patients preferred the DTC remote consultation within 5 days rather than waiting 20 days to consult their RP in person.

**Figure 3.**
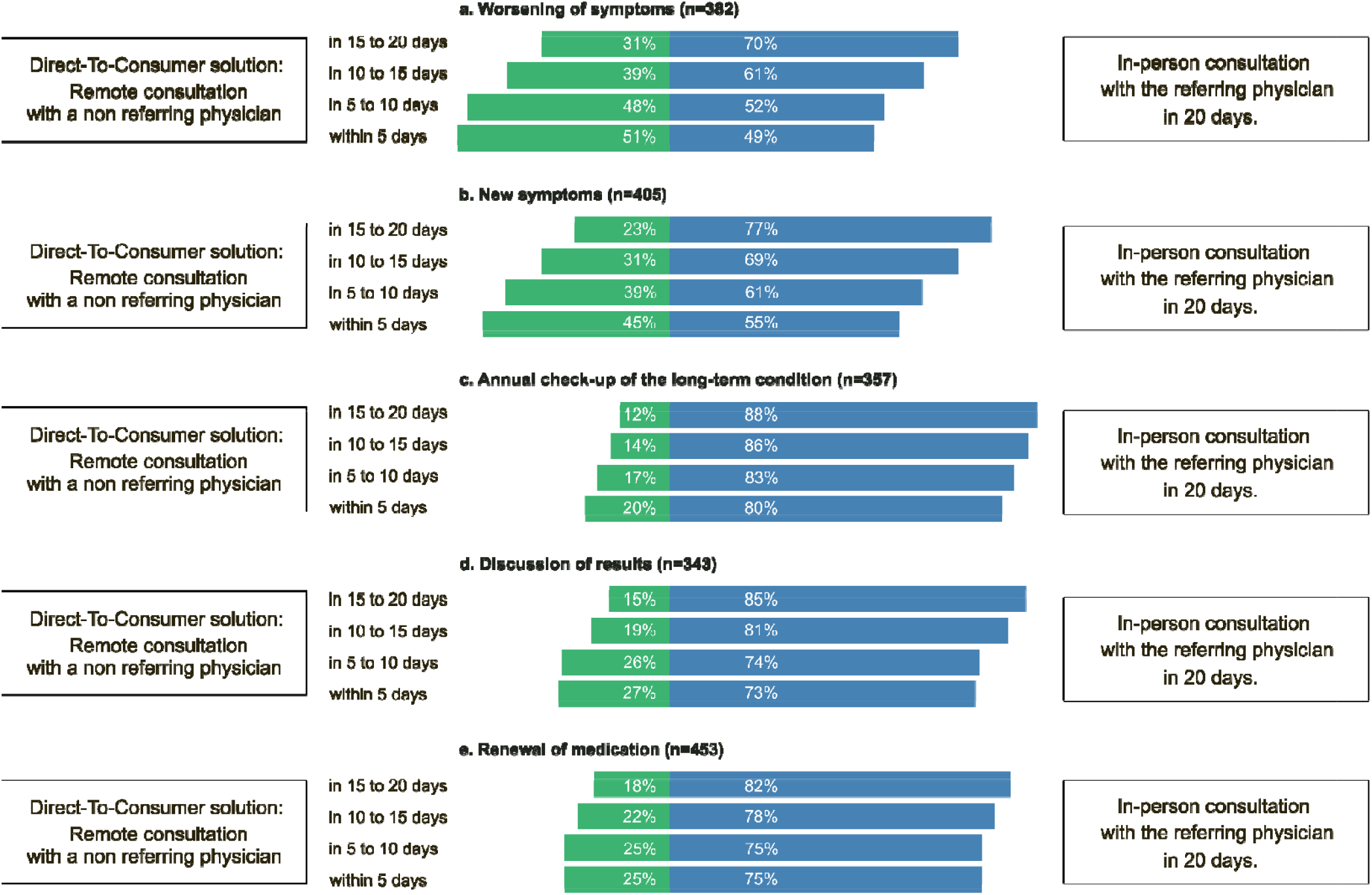
Willingness to trade for a quicker Direct-to-Consumer (DTC) remote consultation with a non-referring physician rather than waiting for an in-person consultation in days with the referring physician (weighted data set, n=1940) Legend: Green bars represent the proportion of patients who prefer the DTC remote consultation with a non-referring physician in the delay mentioned on the left. Blue bars represent the proportion of patients who prefer waiting for the in-person consultation with the referring physician in 20 days. The referring physician is identified by the patients as the physician providing the most regular follow-up of their long-term condition(s). For example, for the worsening of their symptoms (situation a), 31% of patients prefer a DTC remote consultation in 15 to 20 days with a non-referring physician rather than waiting 20 days to consult in person their referring physician. quicker the DTC can be scheduled, the higher this proportion grows: if the DTC remote consultation can be scheduled within 5 days, 51% of patients will opt for the DTC in this situation

In the logistic regression model, clinical situations such as new symptoms (OR 3.16 [2.35-4.26], p<0.001) and worsening symptoms (OR 3.74 [2.77-5.08], p<0.001) were associated to the preference for a quicker DTC remote consultation (with medication renewal as the reference situation) (***Suppl Table 5***). Patients’ characteristics such as the gender male (OR 1.32 [1.04-1.67], p=0.021), the difficulty to consult in person (OR 1.54 [1.22-1.96], p<0.001), and the unavailability of remote modalities with the RP (OR 1.33 [1.09-1.64], p=0.006) were associated with the preference for a quicker DTC remote consultation. Physicians’ characteristics such as the physician specialty (OR 1.79 [1.28-2.50] for general practitioners (GPs), p=0.001), medium or poor listening skills of the RP (OR 1.53 [1.20-1.95], p=0.001) and usual delay < 2 weeks (OR 1.43 [1.12-1.82], p=0.003) were associated with the preference for a quicker DTC remote consultation.

## DISCUSSION

Our nationwide vignette-based study describes the current availability and use of remote care among 1995 adults living with long-term conditions (LTC) in France, their preferences for in-person or remote care with their referring physician, and their willingness to trade for quicker Direct-to-Consumer (DTC) remote consultations depending on the situation. The results were calibrated by weighting to represent the French population of patients with LTC.

We found that for 47% of patients, none of the three remote modalities (video consultation, phone contact, or asynchronous message exchange) was available to interact with their RP. These results are consistent with national statistics, which show that, in France, in 2023, only 4% of consultations were video consultations, a figure that is lower than in other Western countries such as Spain (29%) or Denmark (31%).^26^ Although remote care is expected to improve access for patients with mobility limitations and geographic barriers, its current use in France is skewed toward younger, urban populations, with over half of all video consultations taking place in Paris region and less than 20% in rural areas (representing a third of the population). Another issue regarding the availability and use of remote care is that some patients were unaware of whether remote modalities were offered by their RP.^20^ In 2023, GPs conducted 229 million in-person consultations.^26^ Based on French data, we assumed that our five situations correspond approximately to 58% of GP consultations.^27^ In our study, 37% of patients preferred remote modalities in their assigned situation. We project that at least 49 million in-person GP consultations could be conducted remotely instead of in person (***Suppl Text 3***).

Our findings suggest that up to half of patients would be willing to replace in-person consultations with remote modalities in certain situations, especially when they anticipate that no physical examination or complex decision-making is needed.^10,13,17,20,28^ In these situations, remote care may not only be sufficient but also preferable, especially when it helps reduce the burden of travel, waiting time, and time away from work or family. Synchronous modalities (VC and PC) were preferred over asynchronous message exchanges for all five situations (33% vs. 22% for medication renewal, 30% vs. 6% for worsening symptoms). Our results contrast with a primary care study in Canada, where 82% (out of 13174) of virtual visits were requested via AME rather than synchronously (VC or PC).^15^ One reason that may explain this difference is that secured AME in France was only launched at the national level in 2022. In May 2025, less than one million secure messaging accounts had been opened on this national AME service.^29^ Furthermore, lower digital literacy was associated with a greater preference for in-person consultations, consistent with previous studies.^30^ These results confirm that care delivery has to be adapted to patients’ preferences but also possibilities, without imposing digital tools indiscriminately on all patients.^31^

Our study highlights patients’ willingness to trade for DTC remote consultations when situations felt more urgent, such as the occurrence of new symptoms or worsening of existing symptoms. In these situations, up to 51% of participants preferred quicker DTC remote consultations. This choice results in a loss of continuity of care since the physician providing care is not the RP and has no access to the medical record. This willingness to “trade” continuity for speed may put patients at risk, as continuity of care is associated with better health outcomes.^32^ A recent Canadian study found that patients who underwent a DTC remote consultation were three times more likely to visit the emergency department within 7 days than those who saw their RP.^33^ These findings caution against over-relying on DTC models without ensuring adequate integration into healthcare systems to ensure continuity of care.

This study has several strengths. First, this was a large nationwide study, with participants with diverse LTC recruited from a French e-cohort, which was weighted to approximate the French population of patients living with LTC. Second, we conducted a vignette-based survey, a methodologically robust tool, which allowed us to capture nuanced, context-dependent preferences that might be missed in a single, generic survey about remote care.

Our study also has some limitations. First, our sample does not represent the patients currently using remote care, but rather presents the views of patients with LTC in general. We decided to explore the general willingness of these patients, for whom remote care is likely to be expanded in the future. Second, despite weighting, selection bias is still possible, as many study participants were familiar with digital tools, having been recruited via an e-cohort that required digital literacy. However, patients with lower digital literacy were still represented, with 17% of them scoring lower at the DHCLS. Third, the response rate was relatively low compared to the number of invitations, but this reflects the voluntary nature of participation and the fact that patients were asked to participate in a study outside their usual cohort involvement.

In all, our findings suggest a need for rethinking the organization of follow-up care for patients with LTC. Remote care is underused not because patients reject it, but because it may not be sufficiently offered, accessible, or adapted to their needs. Remote care should not simply be added as a parallel layer to substitute for in-person visits. Instead, it must be thoughtfully integrated into a ‘blended care’ model, where in-person and remote care are complementary, coordinated, and adapted to patients. For example, patients could have direct contact with the physician via asynchronous message exchanges; this contact could be sufficient to address the matter or lead to a consultation scheduled in person or remotely, depending on the patient’s preferences, needs, and possibilities. Providing such care sustainably requires reimagining the organization of physicians and care teams: RP needs time and external help (human and/or technological) to address patients’ asynchronous demands. This approach could reduce unnecessary consultations and preserve in-person care for situations where it brings the most value.^34^

In conclusion, our findings demonstrate that patients with LTC are willing to incorporate remote care into their follow-up, particularly when the modality aligns with the clinical context and personal constraints. Further research on blended care should investigate how to effectively integrate remote care into the healthcare system in a way that maintains patient-centered care and improves patient outcomes, as well as how to reorganize healthcare providers’ time and the healthcare system to make such integration feasible and sustainable.

## Data Availability

All data produced in the present study are available upon reasonable request to the authors

## Author Contributions

Tiphaine Lenfant contributed to conceptualisation, data curation, formal analysis, methodology, project administration, visualisation, writing – original draft, and writing – review and editing.

Elodie Perrodeau contributed to methodology, formal analysis, and writing – review and editing.

Philippe Ravaud contributed to conceptualisation, methodology, supervision, validation, and writing – review and editing.

Viet-Thi Tran contributed to conceptualisation, data curation, formal analysis, methodology, project administration, supervision, validation, visualisation, writing – original draft, and writing – review and editing.

Tiphaine Lenfant and Viet-Thi Tran accessed and verified the underlying data reported in the manuscript. All authors had full access to all the data in the study and accept responsibility for the decision to submit for publication.

## Data sharing statement

The data that support the findings of this study are available from the corresponding author upon reasonable request.

## Use of artificial intelligence

A generative AI tool (ChatGPT, GPT-4, OpenAI) was used solely to improve the language and readability of the manuscript.

