## Supplemental for "Preferences for In-Person Consultations with the Referring Physician versus Remote Care provided by the Referring Physician or another Physician among Patients with Long-Term Conditions: A Vignette-Based Survey"

**Supplementary material**

### Suppl Text 1: Data collection

The first part of the survey gathered:

1) Demographics

- Age (years old)
- Gender (three possible answers: male, female, I prefer not to answer)
- Geographic zone (zip code)
- Highest diploma ("What is the highest diploma you obtained?" divided into 2 levels: “bachelor’s degree or lower” *versus* “higher than a bachelor's degree”)
- Self-perception of financial situation ("How would you describe your current financial situation?", six possible answers were: 1) Very comfortable, 2) Quite comfortable, 3) Balanced, 4) A little tight and I have to be careful with my finances, 5) It's difficult, 6) I am forced to go into debt to get out of it) put into a binary variable with two levels: Yes (answers 1-4) /No (answers 5-6)

2) Health data

- Long-term conditions: "Which condition(s) do you have from this list?" (You can select multiple answers)
  - High blood pressure
  - Diabetes
  - Thyroid disease or other endocrine disease
  - Pulmonary disease (for example, chronic bronchitis or asthma)
  - Heart disease (for example, myocardial infarction, heart failure...)
  - Kidney disease (for example, chronic kidney failure)
  - Gynecological disease (for example, endometriosis)
  - Gastrointestinal disease (for example, ulcer, inflammatory bowel diseases, liver diseases)
  - Neurological disease (for example, stroke, multiple sclerosis, epilepsy)
  - Rheumatological disease (for example, osteoporosis, arthritis, inflammatory rheumatism...)
  - Cancer or hematological disease (for example, lung cancer, prostate cancer, lymphoma, leukemia)
  - Depression, bipolar disorder, chronic anxiety, or other psychiatric disease
  - Vision problems (for example, glaucoma, cataracts...)
  - Dermatological disease (for example, psoriasis, atopic dermatitis)
  - Infectious disease (for example, HIV infection, tuberculosis...)
  - Autoimmune or autoinflammatory disease (for example, lupus, sarcoidosis, Sjögren's syndrome, ANCA-associated vasculitis)
  - Other(s)
- Self-assessment of anxiety linked to their health, self-assessed by the question "Currently, on a scale from 1 to 6, how much do your health problems worry you?" 1: I am not worried about my health at all; 6: I am extremely worried about my health.
- Self-assessment of Self-management, "On a scale from 1 to 6, how do you consider your management of your condition(s) and treatment(s)? 1: I cannot manage them alone at all; 6: I am completely autonomous in managing my illnesses and treatments.
- Digital Health Care Literacy Scale (scale from 3 to 15, 15 being the highest score) put into a binary variable of two levels (lower DHCLS for 3 to 10, higher DHCLS from 11 to 15)

3) Information about their referring physician

- Specialty (General Practitioner, Medical specialty, Surgical Specialty)
- Length of relationship (less than 5 years, 5 to 10 years, more than 10 years)
- Listening skills evaluated by the patient ("How do you evaluate your physician’s ability to listen to you?" with five possible answers 1) Poor 2) Mediocre 3) Average 4) Good 5) Excellent) put into a binary variable Poor to Average (1-3), Good to Excellent (5-6)

4) Information about their usual consultation settings

- Contact modality most frequently used between in-person, video consultation, phone contacts, and asynchronous message exchanges.
- Availability and Frequency of use of remote modalities
  - Video consultation
  - Phone contacts
  - Asynchronous message exchanges
- Time from departure (from home/work) to the start of an in-person consultation, including transportation, parking, inscription, and waiting room, with three possible answers: 1) less than an hour, 2) one to three hours, 3) more than three hours.
- Delay before the next available in-person appointment with the referring physician, assessed by the question "What is the average delay before an in-person consultation with your doctor (for a non-urgent reason)?" with six possible answers: 1) Less than 1 week, 2) Between 1 and 2 weeks, 3) Between 2 weeks and 1 month, 4) Between 1 and 3 months, 5) Between 3 and 6 months, 6) More than 6 months. Divided into a binary variable “less than two weeks” and “two weeks or more”.
- Ability to make personal/professional arrangements to free up time to attend an in-person consultation, assessed by the question "How easy is it for you to free up time to see your doctor?" with four possible answers: 1) It's very easy, I can take a whole day off without any problem. 2) It's easy, I can take half a day off without any problem. 3) It's difficult, I need to make arrangements for family/professional obligations. 4) It's very difficult, almost impossible, to free up time for my appointments. Divided into a binary variable “Able to free up half a day or the entire day” (1-2) and “Difficult to Impossible” (3-4)

### Suppl Text 2: Covariates of the logistic regression models

Covariates were patients’, physicians’, and consultations’ characteristics:

i) *Patients’ characteristics*: age (<60 vs ≥60), gender, economic difficulties (yes/no), highest diploma (up to bachelor’s degree, higher than bachelor’s degree), multimorbidity (1 vs ≥2 LTCs), anxiety score (≤4 vs >4), self-management score (“good or excellent” vs “medium or poor”), DHCLS (≤10, >10/15);
ii) *Physicians’ characteristics*: physician’s listening skills (“good or excellent” vs “medium or poor”), length of relationship (< 5 vs ≥5 years), specialty (general practitioner vs specialist),
iii) *Consultation’s characteristics*: location (at hospital vs in the city), easiness to free up time for the consultation (difficult vs easy), time to consultation (<1 vs ≥1 hour), delay to the next available in-person appointment (<2 weeks, ≥2 weeks), use of remote contact (“not available or never used” vs “used”), *iv)* situation (worsening of symptoms, new symptoms, checkup, renewal, and results).

### Suppl Text 3: Number of in-person GP consultations that could be replaced by remote care in France

General practitioners conducted 229 million in-person consultations in 2023 in France and 5.5 million video consultations.^26^

To approximate the proportion of consultations that align with patients with LTCs and our five clinical scenarios (medication renewal, test result discussion, annual check-up, new symptoms, symptom worsening), we referred to preliminary results of the PaRIS study by the DREES.^27^ In Graph 6 of the report, the categories “Soins courants pour un problème de longue durée” ("routine care for a long-term condition") and “Soins ponctuels pour un problème de longue durée” ("occasional care for a long-term condition") together account for 58% of consultations.

In our study, 37% of patients preferred remote care to in-person consultations in their assigned situation.

We therefore estimated: *229 million of in-person GP consultations × 58% = 132.8 million consultations aligned with our situations*

*132.8 million × 37% = 49.1 million consultations potentially substitutable by remote care*

These are, of course, very rough estimates, meant only to provide an order of magnitude. They likely underestimate the actual potential, as they exclude urgent care and other types of consultations that could also be candidates for remote delivery under certain circumstances.

| **Characteristic** | **RAW**, N = 1,995^1^ | **Worsening symptoms**, N = 393^1^ | **New symptoms**, N = 412^1^ | **Annual checkup**, N = 371^1^ | **Result discussion**, N = 358^1^ | **Medication renewal**, N = 461^1^ |
| --- | --- | --- | --- | --- | --- | --- |
| **Age - years (IQR)** | 52.0 (40.0, 63.0) | 51.0 (38.0, 63.0) | 52.0 (40.8, 64.0) | 53.0 (41.0, 65.0) | 50.0 (38.0, 62.0) | 52.0 (40.0, 62.0) |
| **Gender - no. (%)** |  |  |  |  |  |  |
| Male | 562 (28%) | 127 (32%) | 120 (29%) | 106 (29%) | 88 (25%) | 121 (26%) |
| Female | 1,433 (72%) | 266 (68%) | 292 (71%) | 265 (71%) | 270 (75%) | 340 (74%) |
| **Highest diploma - no. (%)** |  |  |  |  |  |  |
| Higher | 1,209 (61%) | 248 (63%) | 242 (59%) | 215 (58%) | 219 (61%) | 285 (62%) |
| Lower | 786 (39%) | 145 (37%) | 170 (41%) | 156 (42%) | 139 (39%) | 176 (38%) |
| **Financial difficulties — no. (%)** |  |  |  |  |  |  |
| Economical difficulties | 697 (35%) | 137 (35%) | 149 (36%) | 120 (32%) | 123 (34%) | 168 (36%) |
| No difficulties | 1,298 (65%) | 256 (65%) | 263 (64%) | 251 (68%) | 235 (66%) | 293 (64%) |
| **Multimorbidity — no. (%)** | 1,323 (66%) | 241 (61%) | 277 (67%) | 257 (69%) | 232 (65%) | 316 (69%) |
| **Anxiety — no. (%)** |  |  |  |  |  |  |
| 4 or lower | 1,201 (60%) | 239 (61%) | 247 (60%) | 222 (60%) | 216 (60%) | 277 (60%) |
| higher than 4 | 794 (40%) | 154 (39%) | 165 (40%) | 149 (40%) | 142 (40%) | 184 (40%) |
| **Self-management — no. (%)** |  |  |  |  |  |  |
| good or excellent | 1,248 (63%) | 233 (59%) | 253 (61%) | 231 (62%) | 235 (66%) | 296 (64%) |
| medium or poor | 747 (37%) | 160 (41%) | 159 (39%) | 140 (38%) | 123 (34%) | 165 (36%) |
| **DHCLS — no. (%)** |  |  |  |  |  |  |
| higher DHL | 1,729 (87%) | 342 (87%) | 351 (85%) | 317 (85%) | 319 (89%) | 400 (87%) |
| lower DHL | 266 (13%) | 51 (13%) | 61 (15%) | 54 (15%) | 39 (11%) | 61 (13%) |
| **Physician's specialty — no. (%)** |  |  |  |  |  |  |
| general practicioner | 1,259 (63%) | 246 (63%) | 268 (65%) | 212 (57%) | 236 (66%) | 297 (64%) |
| specialist | 735 (37%) | 147 (37%) | 144 (35%) | 158 (43%) | 122 (34%) | 164 (36%) |
| **Length of relationship — no. (%)** |  |  |  |  |  |  |
| 5 years or more | 1,167 (59%) | 234 (60%) | 256 (62%) | 219 (59%) | 191 (54%) | 267 (58%) |
| less than 5 years | 821 (41%) | 158 (40%) | 156 (38%) | 151 (41%) | 163 (46%) | 193 (42%) |
| **Listening skills — no. (%)** |  |  |  |  |  |  |
| good or excellent | 1,553 (78%) | 305 (78%) | 334 (81%) | 286 (78%) | 275 (77%) | 353 (77%) |
| medium or poor | 440 (22%) | 88 (22%) | 78 (19%) | 83 (22%) | 83 (23%) | 108 (23%) |
| **Travel time to consultation — no. (%)** |  |  |  |  |  |  |
| less than an hour | 1,195 (60%) | 245 (63%) | 257 (62%) | 209 (56%) | 215 (60%) | 269 (59%) |
| more than 1 hour | 795 (40%) | 147 (38%) | 155 (38%) | 161 (44%) | 143 (40%) | 189 (41%) |
| **Delay before next consultation — no. (%)** |  |  |  |  |  |  |
| less than 2 weeks | 1,018 (51%) | 204 (52%) | 217 (53%) | 174 (47%) | 191 (54%) | 232 (50%) |
| more than 2 weeks | 970 (49%) | 189 (48%) | 193 (47%) | 194 (53%) | 166 (46%) | 228 (50%) |
| **Easiness to attend in-person — no. (%)** |  |  |  |  |  |  |
| difficult | 564 (28%) | 104 (26%) | 117 (28%) | 96 (26%) | 110 (31%) | 137 (30%) |
| easy | 1,429 (72%) | 289 (74%) | 295 (72%) | 274 (74%) | 248 (69%) | 323 (70%) |
| **Remote interactions — no. (%)** |  |  |  |  |  |  |
| Not available or never used | 857 (43%) | 162 (41%) | 169 (41%) | 167 (45%) | 169 (47%) | 190 (41%) |
| Used | 1,137 (57%) | 231 (59%) | 243 (59%) | 203 (55%) | 189 (53%) | 271 (59%) |
| **Elevated Blood Pressure** | 439 (22%) | 87 (22%) | 102 (25%) | 87 (23%) | 65 (18%) | 98 (21%) |
| **Diabetes** | 203 (10%) | 38 (9.7%) | 39 (9.5%) | 47 (13%) | 34 (9.5%) | 45 (9.8%) |
| **Thyroid disease** | 295 (15%) | 61 (16%) | 49 (12%) | 56 (15%) | 58 (16%) | 71 (15%) |
| **Pulmonary disease** | 221 (11%) | 55 (14%) | 40 (9.7%) | 41 (11%) | 41 (11%) | 44 (9.5%) |
| **Cardiac disease** | 179 (9.0%) | 38 (9.7%) | 34 (8.3%) | 35 (9.4%) | 28 (7.8%) | 44 (9.5%) |
| **Kidney disease** | 108 (5.4%) | 21 (5.3%) | 24 (5.8%) | 25 (6.7%) | 15 (4.2%) | 23 (5.0%) |
| **Gynecological disease** | 373 (19%) | 71 (18%) | 78 (19%) | 67 (18%) | 67 (19%) | 90 (20%) |
| **Digestive disease** | 294 (15%) | 52 (13%) | 54 (13%) | 64 (17%) | 58 (16%) | 66 (14%) |
| **Neurological disease** | 229 (11%) | 37 (9.4%) | 60 (15%) | 44 (12%) | 40 (11%) | 48 (10%) |
| **Rheumatological disease** | 558 (28%) | 100 (25%) | 128 (31%) | 106 (29%) | 107 (30%) | 117 (25%) |
| **Cancer or hematologic** | 161 (8.1%) | 36 (9.2%) | 35 (8.5%) | 28 (7.5%) | 19 (5.3%) | 43 (9.3%) |
| **Mental health disorder** | 530 (27%) | 95 (24%) | 109 (26%) | 90 (24%) | 106 (30%) | 130 (28%) |
| **Ophthalmologic disease** | 193 (9.7%) | 31 (7.9%) | 41 (10.%) | 48 (13%) | 26 (7.3%) | 47 (10%) |
| **Dermatologic disease** | 277 (14%) | 51 (13%) | 52 (13%) | 65 (18%) | 45 (13%) | 64 (14%) |
| **Infectious disease** | 46 (2.3%) | 10 (2.5%) | 13 (3.2%) | 8 (2.2%) | 8 (2.2%) | 7 (1.5%) |
| **Autoimmune disease** | 233 (12%) | 41 (10%) | 47 (11%) | 55 (15%) | 38 (11%) | 52 (11%) |
| **Other** | 525 (26%) | 87 (22%) | 100 (24%) | 113 (30%) | 100 (28%) | 125 (27%) |

### Suppl Table 1: Patients, physicians and consultations characteristics described by clinical situation (n=1995)

| **Characteristics** |  | **In person** | **Remote** | **Odd-ratios (95%CI, p)** |
| --- | --- | --- | --- | --- |
| **Patients’ characteristics** | | | | |
| Age | 60 or older | 420 (67.4) | 203 (32.6) | - |
|  | Younger than 60 | 680 (51.4) | 643 (48.6) | 1.28 (1.00-1.63, p=0.052) |
| Gender | Female | 729 (53.0) | 646 (47.0) | - |
|  | Male | 348 (64.9) | 188 (35.1) | 0.73 (0.57-0.92, p=0.008) |
| Self-perception of financial situation | Difficult | 384 (56.3) | 298 (43.7) | - |
|  | Not difficult | 717 (56.7) | 548 (43.3) | 1.07 (0.86-1.34, p=0.559) |
| Highest diploma | Higher than a bachelor’s degree | 637 (54.0) | 543 (46.0) | - |
|  | Bachelor’s degree or lower | 462 (60.4) | 303 (39.6) | 0.93 (0.75-1.15, p=0.493) |
| Multimorbidity (≥2 LTCs) | No | 370 (56.7) | 282 (43.3) | - |
|  | Yes | 731 (56.4) | 564 (43.6) | 1.22 (0.99-1.52, p=0.066) |
| Self-assessed anxiety level (score from 1 to 6, 6 being the highest level of anxiety) | ≤ 4 | 678 (57.4) | 504 (42.6) | - |
|  | > 4 | 423 (55.3) | 342 (44.7) | 0.98 (0.78-1.21, p=0.823) |
| Self-management (score from 1 to 6, 6 being the best self-management) | Good to excellent (4-6) | 677 (56.2) | 528 (43.8) | - |
|  | Medium to poor (1-3) | 424 (57.1) | 318 (42.9) | 0.88 (0.71-1.09, p=0.240) |
| Digital Health Care Literacy Scale (score from 3 to 15, 15 being the highest) | Higher digital literacy (>10) | 921 (54.3) | 775 (45.7) | - |
|  | Lower digital literacy (≤10) | 180 (71.7) | 71 (28.3) | 0.51 (0.37-0.70, p<0.001) |
| **Referring Physicians’ characteristics** | | | | |
| Listening skills evaluated by the patient | Good to excellent | 882 (58.4) | 628 (41.6) | - |
|  | Medium to poor | 218 (50.0) | 218 (50.0) | 1.35 (1.06-1.73, p=0.016) |
| Length of relationship with the physician | 5 years or more | 662 (58.5) | 469 (41.5) | - |
|  | Less than 5 years | 436 (53.8) | 375 (46.2) | 1.01 (0.82-1.23, p=0.949) |
| Specialty of the physician | General practitioner | 747 (60.3) | 492 (39.7) | - |
|  | Specialist | 354 (50.0) | 354 (50.0) | 1.26 (0.92-1.74, p=0.148) |
| **Consultations’ characteristics** | | | | |
| Location of the consultation | At the hospital | 223 (48.7) | 235 (51.3) | - |
|  | In town | 875 (59.0) | 609 (41.0) | 0.89 (0.64-1.25, p=0.506) |
| Ability to make personal/professional arrangements to free up time to attend an in-person consultation | Difficult to Impossible | 222 (40.1) | 331 (59.9) | - |
|  | Ability to free up half a day or the entire day | 878 (63.0) | 515 (37.0) | 0.47 (0.37-0.60, p<0.001) |
| Time from departure to the start of an in-person consultation (transportation, parking, inscription, waiting room) | Less than an hour | 713 (61.2) | 452 (38.8) | - |
|  | More than 1 hour | 387 (49.7) | 391 (50.3) | 1.20 (0.97-1.50, p=0.095) |
| Delay before the next available in-person consultation | Less than 2 weeks | 626 (63.0) | 368 (37.0) | - |
|  | 2 weeks or more | 471 (49.7) | 476 (50.3) | 1.38 (1.09-1.74, p=0.007) |
| Availability and Use of Remote contacts (VC, PC, AME) | None available or none used | 481 (57.8) | 351 (42.2) | - |
|  | Used | 620 (55.6) | 495 (44.4) | 1.06 (0.86-1.30, p=0.582) |
| Clinical situation | Medication renewal | 176 (39.4) | 271 (60.6) | - |
|  | Annual checkup | 249 (69.6) | 109 (30.4) | 0.27 (0.20-0.37, p<0.001) |
|  | New symptoms | 283 (69.2) | 126 (30.8) | 0.28 (0.21-0.38, p<0.001) |
|  | Worsening of symptoms | 242 (62.9) | 143 (37.1) | 0.39 (0.29-0.53, p<0.001) |
|  | Discussing results | 151 (43.4) | 197 (56.6) | 0.88 (0.65-1.19, p=0.405) |

Suppl Table 2: Factors associated with patients’ preferences for remote modalities with their referring physician for the follow-up of their long-term conditions (logistic regression model). Legend: AME: *Asynchronous Message Exchanges, LTCs: Long Term Conditions, PC: Phone contacts, VC: Video consultation.*

|  |  | **In person** | **Remote** | **Odd-ratios (IC95%, p)** |
| --- | --- | --- | --- | --- |
| Age | 60 or older | 194 (68.8) | 88 (31.2) | - |
|  | Younger than 60 | 276 (51.7) | 258 (48.3) | 1.09 (0.74-1.60, p=0.674) |
| Gender | Female | 302 (52.5) | 273 (47.5) | - |
|  | Male | 168 (69.7) | 73 (30.3) | 0.58 (0.40-0.84, p=0.004) |
| Self-perception of financial situation | Difficult | 171 (56.6) | 131 (43.4) | - |
|  | Not difficult | 299 (58.2) | 215 (41.8) | 0.96 (0.68-1.36, p=0.829) |
| Highest diploma | Higher than a bachelor’s degree | 253 (55.4) | 204 (44.6) | - |
|  | Bachelor’s degree or lower | 217 (60.4) | 142 (39.6) | 0.95 (0.69-1.32, p=0.775) |
| Multimorbidity (≥2 LTCs) | No | 160 (57.3) | 119 (42.7) | - |
|  | Yes | 310 (57.7) | 227 (42.3) | 1.21 (0.86-1.69, p=0.275) |
| Self-assessed anxiety level (score from 1 to 6, 6 being the highest level of anxiety) | ≤ 4 | 288 (57.7) | 211 (42.3) | - |
|  | > 4 | 182 (57.4) | 135 (42.6) | 0.93 (0.65-1.32, p=0.668) |
| Self-management (score from 1 to 6, 6 being the best self-management) | Good to excellent (4-6) | 277 (57.2) | 207 (42.8) | - |
|  | Medium to poor (1-3) | 193 (58.1) | 139 (41.9) | 0.92 (0.66-1.28, p=0.608) |
| Digital Health Care Literacy Scale (score from 3 to 15, 15 being the highest) | Higher digital literacy (>10) | 372 (54.3) | 313 (45.7) | - |
|  | Lower digital literacy (≤10) | 98 (74.8) | 33 (25.2) | 0.43 (0.26-0.68, p<0.001) |
| Listening skills evaluated by the patient | Good to excellent | 346 (61.3) | 218 (38.7) | - |
|  | Medium to poor | 124 (49.2) | 128 (50.8) | 1.32 (0.94-1.85, p=0.109) |
| Length of relationship with the physician | 5 years or more | 294 (61.8) | 182 (38.2) | - |
|  | Less than 5 years | 174 (51.6) | 163 (48.4) | 1.25 (0.92-1.71, p=0.157) |
| Specialty of the physician | General practitioner | 339 (60.1) | 225 (39.9) | - |
|  | Specialist | 131 (52.0) | 121 (48.0) | 1.25 (0.74-2.11, p=0.395) |
| Location of the consultation | At the hospital | 86 (53.1) | 76 (46.9) | - |
|  | In town | 382 (58.6) | 270 (41.4) | 1.25 (0.72-2.17, p=0.438) |
| Ability to make personal/professional arrangements to free up time to attend an in-person consultation | Difficult to Impossible | 92 (38.3) | 148 (61.7) | - |
|  | Ability to free up half a day or the entire day | 378 (65.6) | 198 (34.4) | 0.41 (0.28-0.59, p<0.001) |
| Time from departure to the start of an in-person consultation (transportation, parking, inscription, waiting room) | Less than an hour | 312 (62.0) | 191 (38.0) | - |
|  | More than 1 hour | 157 (50.6) | 153 (49.4) | 1.30 (0.93-1.82, p=0.119) |
| Delay before the next available in-person consultation | Less than 2 weeks | 276 (63.0) | 162 (37.0) | - |
|  | 2 weeks or more | 192 (51.2) | 183 (48.8) | 1.34 (0.92-1.94, p=0.124) |
| Clinical situation | Medication renewal | 82 (44.3) | 103 (55.7) | - |
|  | Annual checkup | 102 (64.6) | 56 (35.4) | 0.46 (0.28-0.73, p=0.001) |
|  | New symptoms | 117 (72.7) | 44 (27.3) | 0.30 (0.18-0.48, p<0.001) |
|  | Worsening of symptoms | 90 (59.6) | 61 (40.4) | 0.56 (0.35-0.89, p=0.015) |
|  | Discussing results | 79 (49.1) | 82 (50.9) | 0.90 (0.57-1.42, p=0.636) |

Suppl Table 3: Factors associated with the preference for a remote modality over an in-person consultation with the referring physician among unserved patients (patients with no access to any of the three remote modalities to interact with their physician), logistic regression model. *Legend:* *LTCs: Long-term Conditions.*

|  | **Raw (n=1940)** | | **Weighted (n=1940)** | | **Worsening (n=382)** | **New (n=405)** | **Checkup (n=357)** | **Renewal (n=453)** | **Results (n=343)** |
| --- | --- | --- | --- | --- | --- | --- | --- | --- | --- |
|  | n | % | n | % | % | % | % | % | % |
| ***"Would you like to see an unknown physician sooner remotely (Direct-To-Consumer) rather than you referring physician in 20 days in person ?"*** | | | | | | | | | |
| It depends on the delay | **358** | 18,5% | **359** | 18,5% | 25,6% | 28,9% | 11,0% | 11,5% | 14,5% |
| Yes, I would opt for this DTC appointment | 349 | 18,0% | 342 | 17,6% | 26,6% | 20,0% | 10,7% | 16,4% | 14,0% |
| No, I prefer my physician in person in 20 days | 1233 | 63,6% | 1239 | 63,8% | 47,8% | 51,1% | 78,3% | 72,1% | 71,5% |
| total answers | 1940 |  | 1940 |  |  |  |  |  |  |
| ***For patients answering they would consider seeing an unknown physician depending on the delay*** | | | | | | | | | |
| ***If the DTC appointment is in 15-20 days ?*** |  |  |  |  |  |  |  |  |  |
| Total patients asked | 358 | 100% | 359 | 100,0% |  |  |  |  |  |
| Yes, I would opt for this DTC appointment | 33 | 9,2% | 41 | 11,4% | 16,0% | 9,8% | 11,3% | 16,0% | 3,4% |
| No, I prefer my physician in person in 20 days | **322** | 89,9% | **316** | 88,0% | 84,0% | 90,2% | 88,7% | 84,0% | 96,6% |
| *missing* | *3* | *0,8%* | *2* | *0,6%* |  |  |  |  |  |
| ***If the DTC appointment is in 10-15 days ?*** |  |  |  |  |  |  |  |  |  |
| Total patients asked | 322 | 100% | 316 | 100,0% |  |  |  |  |  |
| Yes, I would opt for this DTC appointment | 101 | 31,4% | 103 | 32,6% | 39,3% | 30,5% | 21,7% | 38,7% | 31,9% |
| No, I prefer my physician in person in 20 days | **218** | 67,7% | **210** | 66,5% | 60,7% | 69,5% | 78,3% | 61,3% | 68,1% |
| *missing* | *3* | *0,9%* | *3* | *0,9%* |  |  |  |  |  |
| ***If the DTC appointment is in 5-10 days ?*** |  |  |  |  |  |  |  |  |  |
| Total patients asked | 218 | 100% | 210 | 100,0% |  |  |  |  |  |
| Yes, I would opt for this DTC appointment | 126 | 57,8% | 118 | 56,2% | 70,8% | 47,5% | 42,1% | 53,9% | 70,8% |
| No, I prefer my physician in person in 20 days | **92** | 42,2% | **92** | 43,8% | 29,2% | 52,5% | 57,9% | 46,1% | 29,2% |
| *missing* | *0* | *0,0%* | 0 | 0,0% |  |  |  |  |  |
| ***If the DTC appointment is in the next 5 days ?*** |  |  |  |  |  |  |  |  |  |
| Total patients asked | 92 | 100% | 92 | 100,0% |  |  |  |  |  |
| Yes, I would opt for this DTC appointment | 52 | 56,5% | 45 | 48,9% | 65,8% | 56,9% | 68,8% | 2,8% | 33,8% |
| No, I prefer my physician in person in 20 days | **38** | 41,3% | **43** | 46,7% | 34,2% | 43,1% | 31,2% | 97,2% | 66,2% |
| *missing* | *2* | *2,2%* | *4* | *4,3%* |  |  |  |  |  |
| ***Compared to in person with referring in 20 days, I prefer the unknown physician remotely if the appointment is in …*** | | | | | | | | | |
|  |  |  | **Weighted (n=1940)** | | **Worsening (n=382)** | **New (n=405)** | **Checkup (n=357)** | **Renewal (n=453)** | **Results (n=343)** |
|  |  |  |  | % | % | % | % | % | % |
| DTC in a shorter delay without precision |  | shorter |  | 17,6% | 26,6% | 20,0% | 10,7% | 16,4% | 14,0% |
| DTC in a shorter delay + 15-20 days |  | 15-20 d |  | 19,7% | 30,7% | 22,8% | 11,9% | 18,2% | 14,5% |
| DTC in a shorter delay + 15-20 d + 10-15 days |  | 10-15 d |  | 25,0% | 39,1% | 30,8% | 14,1% | 22,0% | 18,9% |
| DTC in a shorter delay + 15-20 d + 10-15 d + 5-10 days |  | 5-10 d |  | 31,1% | 48,4% | 39,4% | 17,3% | 25,2% | 25,7% |
| DTC in a shorter delay + 15-20 d + 10-15 d + 5-10 d + < 5 d |  | < 5 d |  | 33,4% | 50,9% | 44,8% | 20,3% | 25,2% | 26,6% |

### Suppl Table 4: Patients’ willingness to trade for a quicker DTC remote consultation with a non-referring physician rather than waiting 20 days to consult their referring physician in person, depending on the delay before the DTC and the clinical situation (weighted data set, n=1940).

|  |  | **Wait 20 days for an in-person consultation with the referring physician** | **DTC solution in 20 days or less (remote consultation with non-referring physician)** | **Odd-ratios (95%CI , p)** |
| --- | --- | --- | --- | --- |
| **Patients’ characteristics** | | | | |
| Age | 60 or older | 413 (64.3) | 229 (35.7) | - |
|  | Younger than 60 | 837 (62.6) | 499 (37.4) | 0.99 (0.77-1.27, p=0.922) |
| Gender | Female | 909 (65.3) | 484 (34.7) | - |
|  | Male | 325 (59.0) | 226 (41.0) | 1.32 (1.04-1.67, p=0.021) |
| Self-perception of financial situation | Difficult | 434 (63.0) | 255 (37.0) | - |
|  | Not difficult | 816 (63.2) | 475 (36.8) | 0.94 (0.75-1.18, p=0.587) |
| Highest diploma | Higher than a bachelor’s degree | 746 (62.3) | 452 (37.7) | - |
|  | Bachelor’s degree or lower | 503 (64.5) | 277 (35.5) | 0.87 (0.70-1.08, p=0.219) |
| Multimorbidity (≥2 LTCs) | No | 403 (60.5) | 263 (39.5) | - |
|  | Yes | 847 (64.5) | 467 (35.5) | 0.87 (0.70-1.08, p=0.204) |
| Self-assessed anxiety level (score from 1 to 6, 6 being the highest level of anxiety) | ≤ 4 | 747 (62.4) | 451 (37.6) | - |
|  | > 4 | 503 (64.3) | 279 (35.7) | 0.96 (0.77-1.20, p=0.733) |
| Self-management (score from 1 to 6, 6 being the best self-management) | Good to excellent (4-6) | 780 (63.4) | 450 (36.6) | - |
|  | Medium to poor (1-3) | 470 (62.7) | 280 (37.3) | 0.93 (0.75-1.16, p=0.532) |
| Digital Health Care Literacy Scale (score from 3 to 15, 15 being the highest) | Higher digital literacy (>10) | 1074 (62.4) | 648 (37.6) | - |
|  | Lower digital literacy (≤10) | 176 (68.2) | 82 (31.8) | 0.81 (0.59-1.11, p=0.195) |
| **Referring Physicians’ characteristics** | | | | |
| Listening skills evaluated by the patient | Good to excellent | 1008 (65.4) | 534 (34.6) | - |
|  | Medium to poor | 241 (55.1) | 196 (44.9) | 1.53 (1.20-1.95, p=0.001) |
| Length of relationship with the physician | 5 years or more | 736 (63.9) | 415 (36.1) | - |
|  | Less than 5 years | 509 (61.8) | 315 (38.2) | 1.17 (0.95-1.44, p=0.134) |
| Specialty of the physician | Specialist | 511 (70.6) | 213 (29.4) | - |
|  | General practitioner | 739 (58.8) | 517 (41.2) | 1.79 (1.28-2.50, p=0.001) |
| **Consultations’ characteristics** | | | | |
| Location of the consultation | At the hospital | 318 (68.1) | 149 (31.9) | - |
|  | In town | 930 (61.7) | 577 (38.3) | 0.72 (0.50-1.02, p=0.068) |
| Ability to make personal/professional arrangements to free up time to attend an in-person consultation | Ability to free up half a day or the entire day | 934 (65.5) | 491 (34.5) | - |
|  | Difficult to Impossible | 316 (57.0) | 238 (43.0) | 1.54 (1.22-1.96, p<0.001) |
| Time from departure to the start of an in-person consultation (transportation, parking, inscription, waiting room) | Less than an hour | 745 (62.4) | 448 (37.6) | - |
|  | More than 1 hour | 504 (64.4) | 279 (35.6) | 1.08 (0.87-1.36, p=0.483) |
| Delay before the next available in-person consultation | 2 weeks or more | 654 (68.2) | 305 (31.8) | - |
|  | Less than 2 weeks | 592 (58.3) | 423 (41.7) | 1.43 (1.12-1.82, p=0.003) |
| Availability and Use of Remote contacts (VC, PC, AME) | Used | 751 (66.2) | 383 (33.8) | - |
|  | None available or none used (unserved) | 499 (59.0) | 347 (41.0) | 1.33 (1.09-1.64, p=0.006) |
| Clinical situation | Medication renewal | 333 (72.7) | 125 (27.3) | - |
|  | Annual checkup | 289 (79.4) | 75 (20.6) | 0.75 (0.54-1.06, p=0.103) |
|  | New symptoms | 198 (47.6) | 218 (52.4) | 3.16 (2.35-4.26, p<0.001) |
|  | Worsening of symptoms | 171 (43.3) | 224 (56.7) | 3.74 (2.77-5.08, p<0.001) |
|  | Discussing results | 259 (74.6) | 88 (25.4) | 0.89 (0.64-1.23, p=0.485) |

Suppl Table 5: Factors associated with patients’ preferences for a quicker Direct-To-Consumer remote consultation with a non-referring physician rather than an in-person consultation with their referring physician in 20 days, (logistic regression model). *Legend: AME: Asynchronous Message Exchanges, DTC: Direct-To-Consumer (remote consultation with a non-referring physician), LTCs: Long Term Conditions, PC: Phone contacts, VC: Video consultation*

### Suppl Figure 1: Flow chart

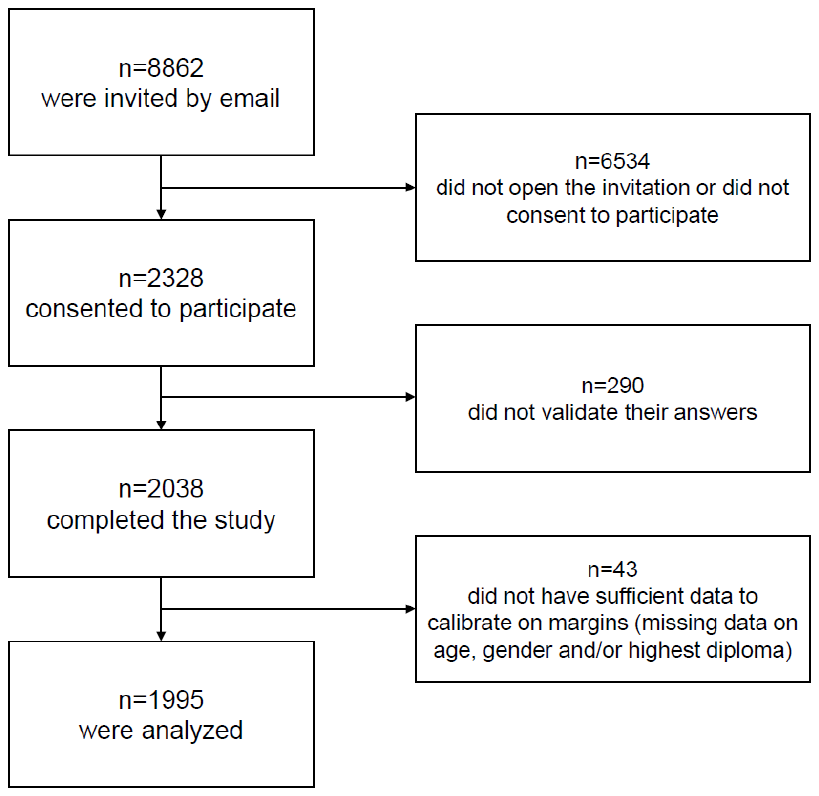

Suppl Figure 2: Preferences for in-person consultation or remote modalities with the referring physician, depending on the clinical situation (weighted data set, n=1908) of all patients (panel a) and of unserved patients (panel b). *Legend: unserved patients were patients who had no access to or never used any remote modality with their referring physician.*

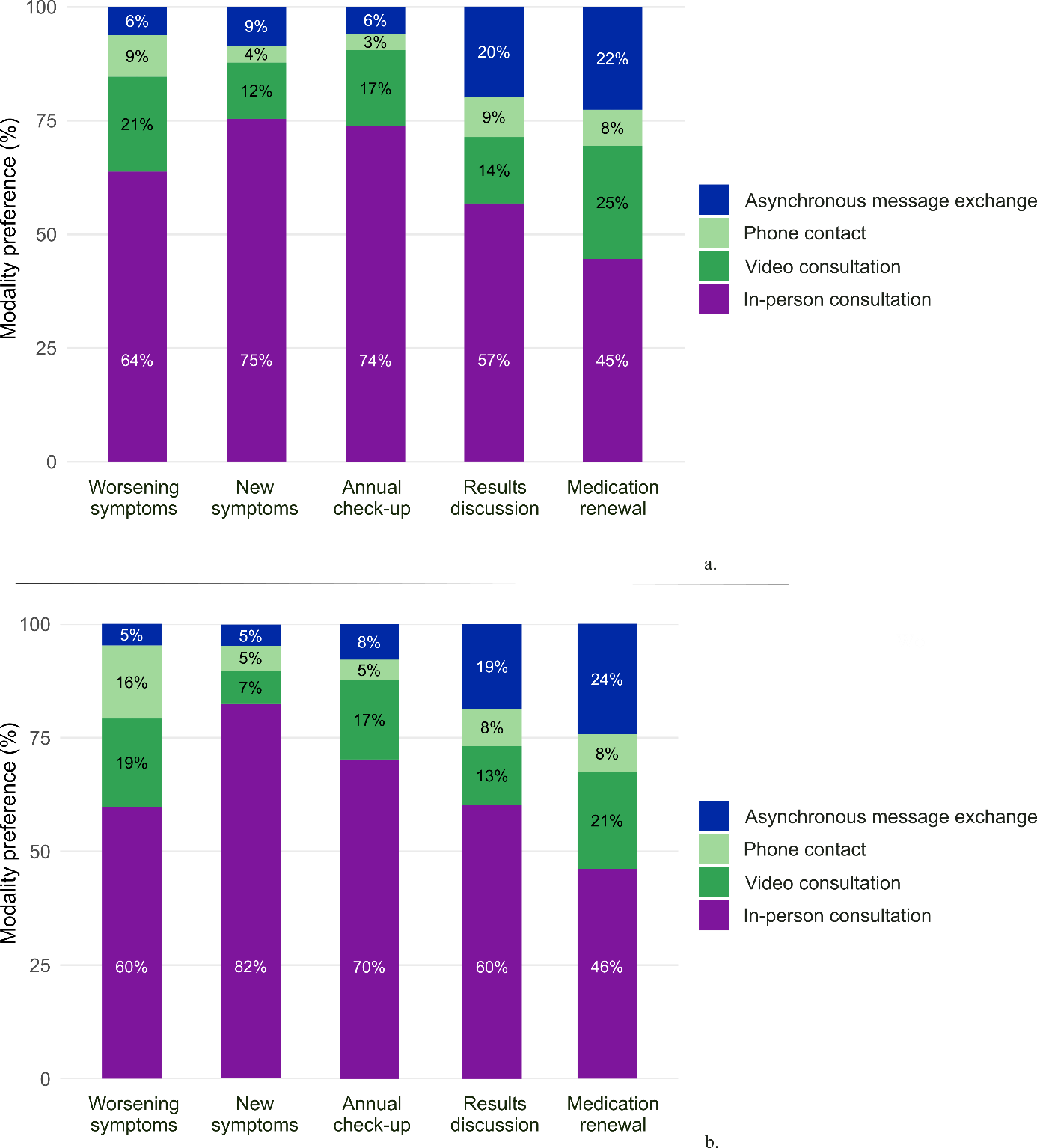
